# Assessment of compliance with quality laboratory standards in medical laboratories within the Kumasi Metropolis, Ghana

**DOI:** 10.1101/2025.10.13.25337932

**Authors:** Elizabeth Sorvor, Mawuli Dzodzomenyo, Justice Nonvignon, Genevieve Cecilia Aryeetey

## Abstract

**Background:** In resource-limited settings, compliance with laboratory standards remains a significant challenge, resulting in delays and inaccurate results that can compromise emergency responses, clinical decision-making, and patient outcomes. In Ghana, medical laboratories are a vital component of the health system, but there has been limited evidence on how well they meet international quality standards. This study assessed compliance with laboratory standards by medical laboratories in the Kumasi Metropolis of Ghana.

**Methods:** A descriptive cross-sectional survey of forty-three (43) laboratory facilities operating within healthcare facilities and private standalone laboratory facilities in the Kumasi metropolis was conducted from January to December 2021. The WHO SLIPTA checklist, which specifies the requirements for the quality and competency of laboratories, was used to assess compliance with laboratory standards. Descriptive analysis and the Kruskal-Wallis test were conducted to examine the differences in overall compliance scores across the types of health facilities.

**Results:** The median compliance score was 61(interquartile range 53-66), the minimum score was 49, and the maximum score was 104. Tertiary-level facility laboratories performed better (93) compared with secondary (66) and primary (64.8) level facilities. Similarly, faith-based facilities (CHAG) performed better (80.8) than private hospitals (60.1) and private standalone laboratories (54.3). There were significant differences in the overall median compliance score (p=0.00) among the four (4) categories of health facilities.

**Conclusion:** The degree of compliance with quality laboratory standards was low across all facility types. Medical laboratories not meeting basic quality standards could significantly impact the accuracy, reliability, and timeliness of test results.

## Introduction

Medical laboratory services have evolved to become an integral part of healthcare delivery systems worldwide [1,2]. The role of the medical laboratory is recognized as critical in the general improvement of countries’ health systems [3]. Laboratory test results are widely used in clinical and public health settings, and health outcomes depend on the accuracy of the testing. The assertion that “ seventy percent (70%) of clinical decisions depend on laboratory data” has become increasingly undeniable [4].

According to Mahatma Gandhi, “A correct diagnosis is three-fourths the remedy”. It has been shown that the medical laboratory is among the top three functions for attaining improved key performance indicators(KPIs), the most prominent in health informatics infrastructure, and an unexploited source for greater value in healthcare delivery [5]. Concerns about the quality of laboratory testing have led to the establishment of regulations, standard guidelines, and quality improvement programs in the medical laboratory system to reduce errors associated with testing [6].

In Ghana, there are about 800 public sector laboratories [7], however, only five (5) of the public sector laboratories, which conduct the majority of patient testing, have recently received accreditation by international standards [8,9]. The healthcare systems in Ghana are plagued with varying degrees of medical laboratory diagnostic inefficiencies, and in some instances, laboratories are not equipped to respond to local health needs. Studies on medical laboratories in Ghana are limited, however, a recent study conducted on gaps in laboratory quality management systems focused on only six(6) laboratories [10], making it difficult to conclude on the overall quality of medical laboratory services. This study aims to evaluate compliance with quality laboratory standards in one of Ghana’s largest metropolitan areas in the Ashanti region.

## Methods

### Study design and setting

The study employed a descriptive cross-sectional design. The study was conducted in the Kumasi Metropolis of the Ashanti region of Ghana. The region’s unique central position makes it accessible from all corners of the country. Kumasi is the second-largest city in the country and the administrative capital of the Ashanti region. Kumasi has the highest number of healthcare facilities, representing 38% of the total number of facilities in the Ashanti region. There are about two hundred and fifteen (215) health facilities in the metropolis, made up of hospitals, health centres, standalone laboratories, and clinics. The majority (87%) of health facilities in the metropolis are privately owned. For this study, only hospitals that provide medical laboratory services were recruited.

### Study population and sample

The study population was healthcare facilities that provide medical laboratory services. A Census of all medical laboratories in healthcare facilities in the Kumasi metropolis was conducted to assess compliance with quality laboratory standards.

### Instrument

The WHO SLIPTA (Stepwise Laboratory Quality Improvement Process Towards Accreditation) checklist was adapted to measure compliance with laboratory standards. The elements of this checklist are based on ISO15189:2012 and CLSI guidelines on Quality Management System: A Model for Laboratory Services[11]. It is a requirement for quality and competence for laboratories to demonstrate the quality and reliability of their services. The checklist assessment enables the determination of whether a laboratory is providing accurate and reliable results, it is well-managed and adhering to good laboratory practices, and identifies areas for improvement[11].

The checklist contains 12 main sections, with 117 questions for a total of 275 scores. Each item is awarded a point value of 2, 3, or 5 points based on relative importance and complexity. Scores of 0-150, 151-177,178-205,206-232,233-260 and 261-275, ultimately translate to 0 to 5 stars, respectively (Table 1). However, for this study, compliance rates were categorised as low (150 and below), moderate (between 151 and 232) and high (above 232).

**Table 1:**
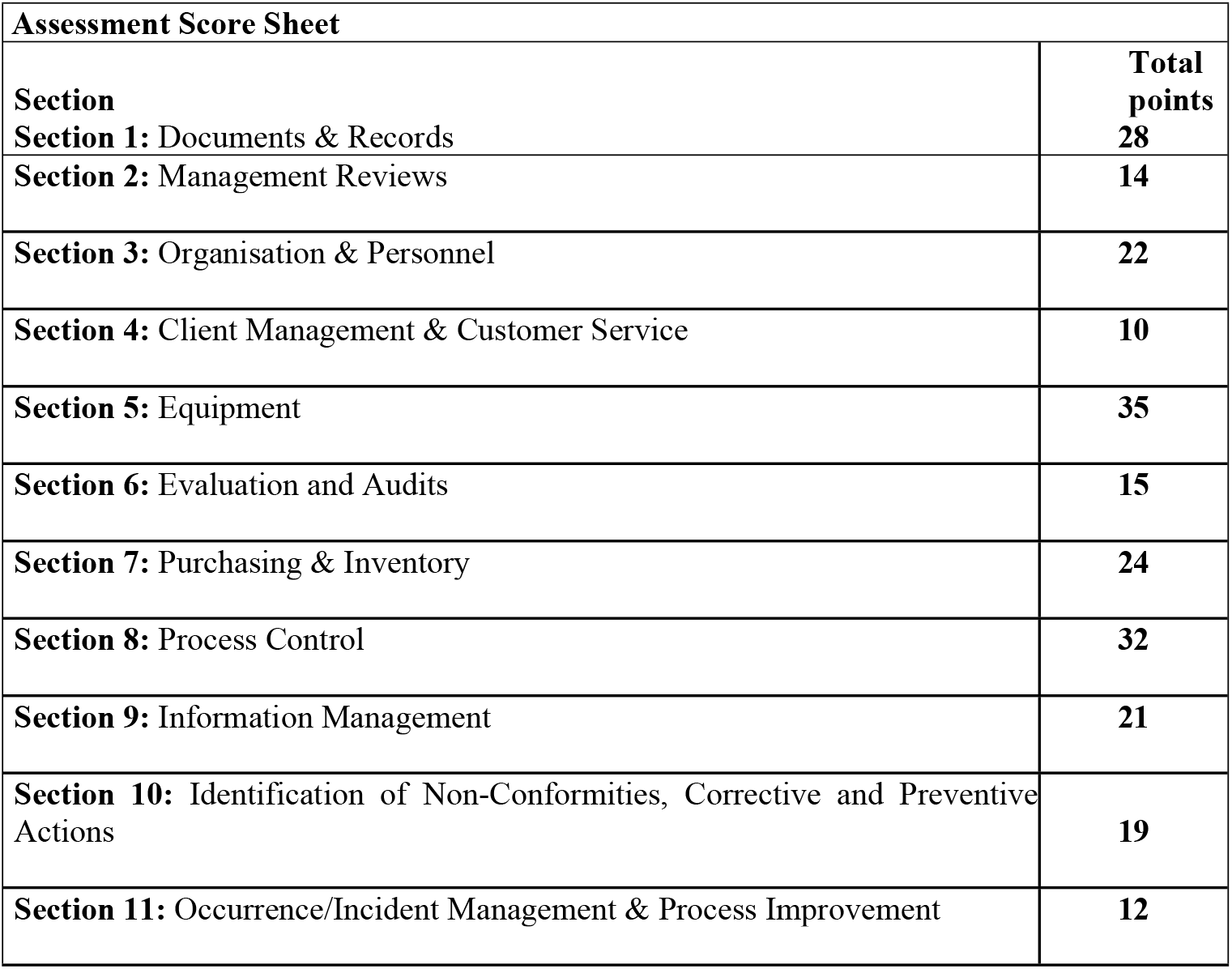
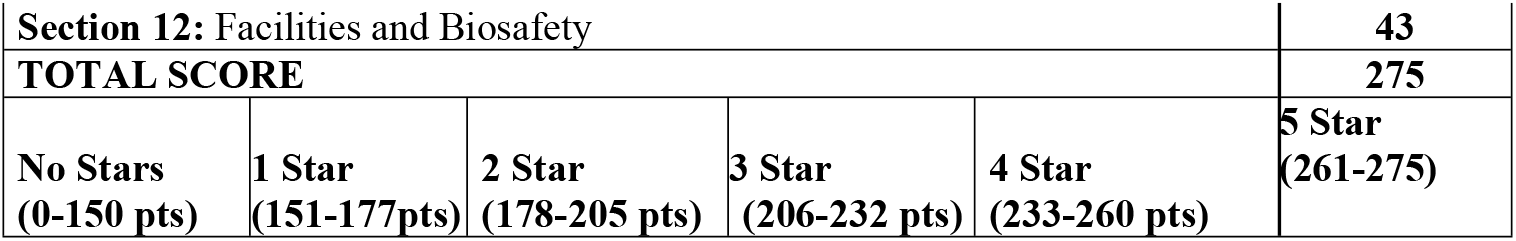
Items Included in the Questionnaire and Score Sheets.

| **Assessment Score Sheet** |  |
| --- | --- |
| **Section** | **Total points** |
| **Section 1:** Documents & Records | **28** |
| **Section 2:** Management Reviews | **14** |
| **Section 3:** Organisation & Personnel | **22** |
| **Section 4:** Client Management & Customer Service | **10** |
| **Section 5:** Equipment | **35** |
| **Section 6:** Evaluation and Audits | **15** |
| **Section 7:** Purchasing & Inventory | **24** |
| **Section 8:** Process Control | **32** |
| **Section 9:** Information Management | **21** |
| **Section 10:** Identification of Non-Conformities, Corrective and Preventive Actions | **19** |
| **Section 11:** Occurrence/Incident Management & Process Improvement | **12** |

|  |  |  |  |  |  |
| --- | --- | --- | --- | --- | --- |
| **Section 12: Facilities and Biosafety** |  |  |  |  | **43** |
| **TOTAL SCORE** |  |  |  |  | **275** |
| **No Stars**<br>**(0-150 pts)** | **1 Star**<br>**(151-177pts)** | **2 Star**<br>**(178-205 pts)** | **3 Star**<br>**(206-232 pts)** | **4 Star**<br>**(233-260 pts)** | **5 Star**<br>**(261-275)** |

### Data collection

The laboratory assessment was conducted from January 2021 to December 2021. Data was collected based on the requirements from the checklist, using a structured questionnaire, observation and document review. The checklist was administered to each laboratory by reviewing laboratory documents to verify that the laboratory quality manual, policies, Standard Operating Procedures (SOPs), and other manuals are complete, current, accurate, and annually reviewed. Additionally, laboratory records such as equipment maintenance records, audit trails, incident reports, logs, personnel files, Internal Quality Control (IQC) records and External Quality Assurance records (EQA) were reviewed. Furthermore, laboratory operations were observed to ensure laboratory testing follows written policies and procedures in pre-analytic, analytic and post-analytic phases of laboratory testing, laboratory procedures are appropriate for the testing performed, deficiencies and nonconformities identified are adequately investigated and resolved within the established timeframe [11].

### Data analysis

Data were imputed into Microsoft Excel sheets, coded, and analysed using Stata version 14 software (Stata Corp LLC). The data’s normal distribution was assessed using the Kolmogorov– Smirnov test, which rejected the normality hypothesis (P <.001). Consequently, nonparametric tests were performed on the data. Descriptive analysis and the Kruskal-Wallis Test were conducted to examine the differences in median compliance scores according to the types of health facilities.

### Ethical consideration

This study is part of the PhD research work of the first author, and received ethical approval from the Ghana Health Service Ethics Review Committee (Reference #: GHS-ERC: 014/11/20) and Komfo Anokye Teaching Hospital Institutional Review Board (IRB) (Reference #: KATH IRB/AP/155/20). However, this part of the study did not involve human participants or identifiable personal data.

## Results

### Characteristics of health facilities and human resource profile

A total of forty-three (43) medical laboratories were assessed. The majority (72%) of laboratories were privately owned health facilities (private hospitals and laboratories), and the least (4%) were government-owned tertiary and secondary hospitals (Table 2). There was a 40% and 67% deficit of medical laboratory scientists in government-owned Tertiary and Secondary health facilities, respectively. However, there was a surplus of medical laboratory scientists in primary (14.3%), private hospitals (209%) and CHAG (53.8%) facilities (Table 3).

**Table 2:**
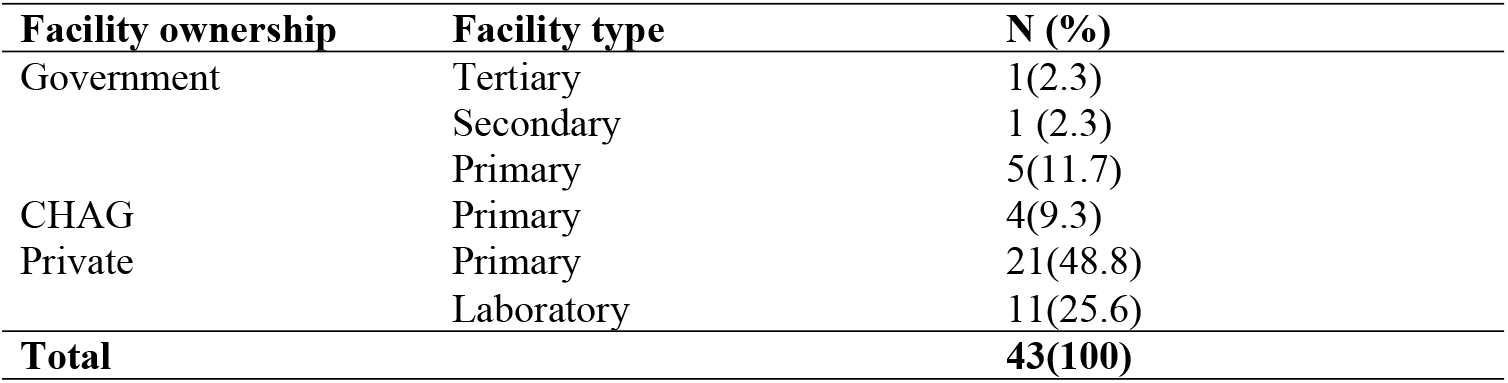
Background Characteristics of Health Facilities.

| Facility ownership | Facility type | N (%) |
| --- | --- | --- |
| Government | Tertiary | 1(2.3) |
|  | Secondary | 1 (2.3) |
|  | Primary | 5(11.7) |
| CHAG | Primary | 4(9.3) |
| Private | Primary | 21(48.8) |
|  | Laboratory | 11(25.6) |
| **Total** |  | **43(100)** |

**Table 3:**
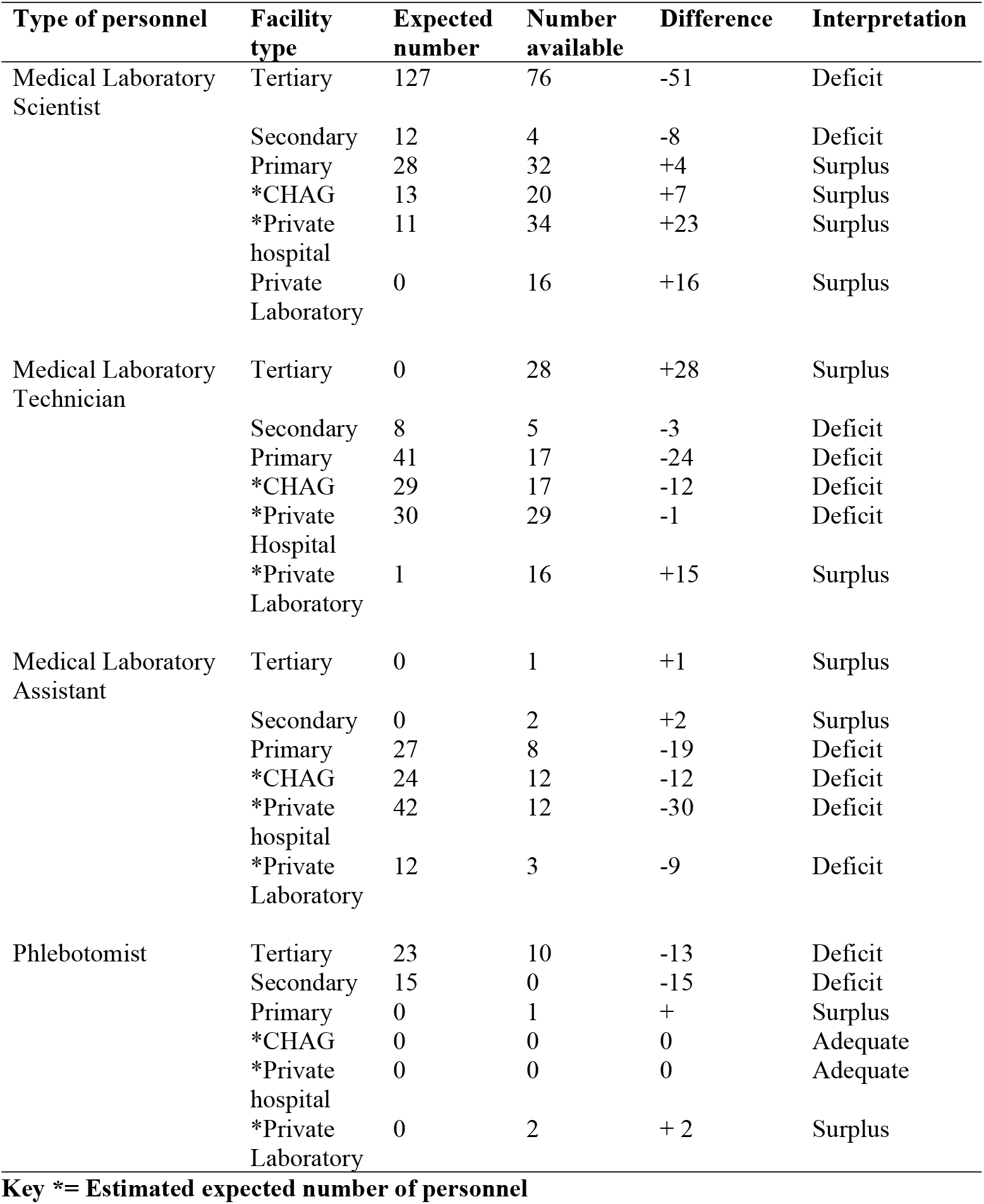
Human Resource Profile of Health Facilities.

### Laboratory compliance scores by facility type

The median compliance score was 61(interquartile range 53-66), the minimum score was 49, and the maximum score was 104. Tertiary-level facility laboratories performed better (93) compared with secondary (66) and Primary (64.8) level facilities. Similarly, faith-based organisations (CHAG) performed better (80.8) than Private hospitals (60.1) and Private Standalone laboratories (54.3) (Table 4). All laboratories performed poorly on the twelve quality system essentials across different facilities (Table 5). The top three performing quality system essentials were purchasing and inventory (11), equipment (8) and facilities and biosafety (14). However, all facilities performed poorly in the following quality indicators: document and records (2), management review and responsibility (0), Client Management and Customer Service (3), Evaluation and Audits (0), Identification of Nonconformities, Corrective Action and Preventive Actions (0), and Occurrence Management (0).

**Table 4:** Overall Compliance Score by Each Laboratory.

| Facility | Ownership | Compliance score |
| --- | --- | --- |
| CHG01 | CHAG | 84 |
| CHG02 | CHAG | 66 |
| CHG03 | CHAG | 104 |
| CHG04 | CHAG | 69 |
| G001 | Government | 63 |
| G002 | Government | 67 |
| G003 | Government | 66 |
| G004 | Government | 64 |
| G005 | Government | 62 |
| G006 | Government | 68 |
| G007 | Government | 93 |
| P001 | Private | 72 |
| P002 | Private | 62 |
| P003 | Private | 62 |
| P004 | Private | 80 |
| P005 | Private | 74 |
| P006 | Private | 58 |
| P007 | Private | 59 |
| P008 | Private | 68 |
| P009 | Private | 53 |
| P010 | Private | 59 |
| P011 | Private | 57 |
| P012 | Private | 53 |
| P013 | Private | 61 |
| P014 | Private | 55 |
| P015 | Private | 55 |
| P016 | Private | 56 |
| P017 | Private | 53 |
| P018 | Private | 54 |
| P019 | Private | 48 |
| P021 | Private | 64 |
| P022 | Private | 57 |
| S001 | Standalone | 63 |
| S002 | Standalone | 54 |
| S003 | Standalone | 50 |
| S004 | Standalone | 50 |
| S005 | Standalone | 49 |
| S006 | Standalone | 49 |
| S007 | Standalone | 64 |
| S008 | Standalone | 49 |
| S009 | Standalone | 52 |
| S010 | Standalone | 66 |
| S011 | Standalone | 51 |

**Table 5:** Laboratory Performance in the Quality System Essentials by Facility Type.

| Quality system essentials | Government |  |  | Govt. | CHAG | Private |  | Expected scores |
| --- | --- | --- | --- | --- | --- | --- | --- | --- |
|  | Tertiary | Secondary | Primary |  |  | Hospital | Laboratory |  |
| Document & Records | 4.0 | 2.0 | 2.0 | 3.0 | 2.8 | 2.2 | 2.0 | 28 |
| Management Review & Responsibility | 0 | 0 | 0 | 0.0 | 0.3 | 0 | 0 | 14 |
| Organization & Personnel | 11.0 | 10.0 | 8.2 | 9.7 | 8.5 | 5.8 | 4.9 | 22 |
| Client Management and Customer Service | 2.0 | 3.0 | 2.0 | 2.3 | 3.5 | 2.7 | 2.3 | 10 |
| Equipment | 15.0 | 7.0 | 9.2 | 10.4 | 14.3 | 9.0 | 7.4 | 35 |
| Evaluation & Audits | 0 | 0 | 0 | 0.0 | 0 | 0 | 0 | 15 |
| Purchasing & Inventory | 11.0 | 10.0 | 13.0 | 11.3 | 13.8 | 11.9 | 10.5 | 24 |
| Process control | 15.0 | 9.0 | 9.0 | 11.0 | 8.5 | 7.5 | 6.5 | 32 |
| Information management | 10.0 | 8.0 | 7.6 | 8.5 | 11.0 | 7.7 | 6.8 | 21 |
| Identification of NCCPA* | 1.0 | 0 | 0 | 0.3 | 0.3 | 0 | 0 | 19 |
| Occurrence management | 4.0 | 0 | 0 | 1.3 | 1.3 | 0.0 | 0 | 12 |
| Facilities & Biosafety | 20.0 | 17.0 | 13.8 | 16.9 | 16.8 | 13.2 | 13.8 | 43 |
| **Overall compliance score** | **93.0** | **66.0** | **64.8** | **74.7** | **81.1** | **60.1** | **54.2** | **275** |

**Table 6:**
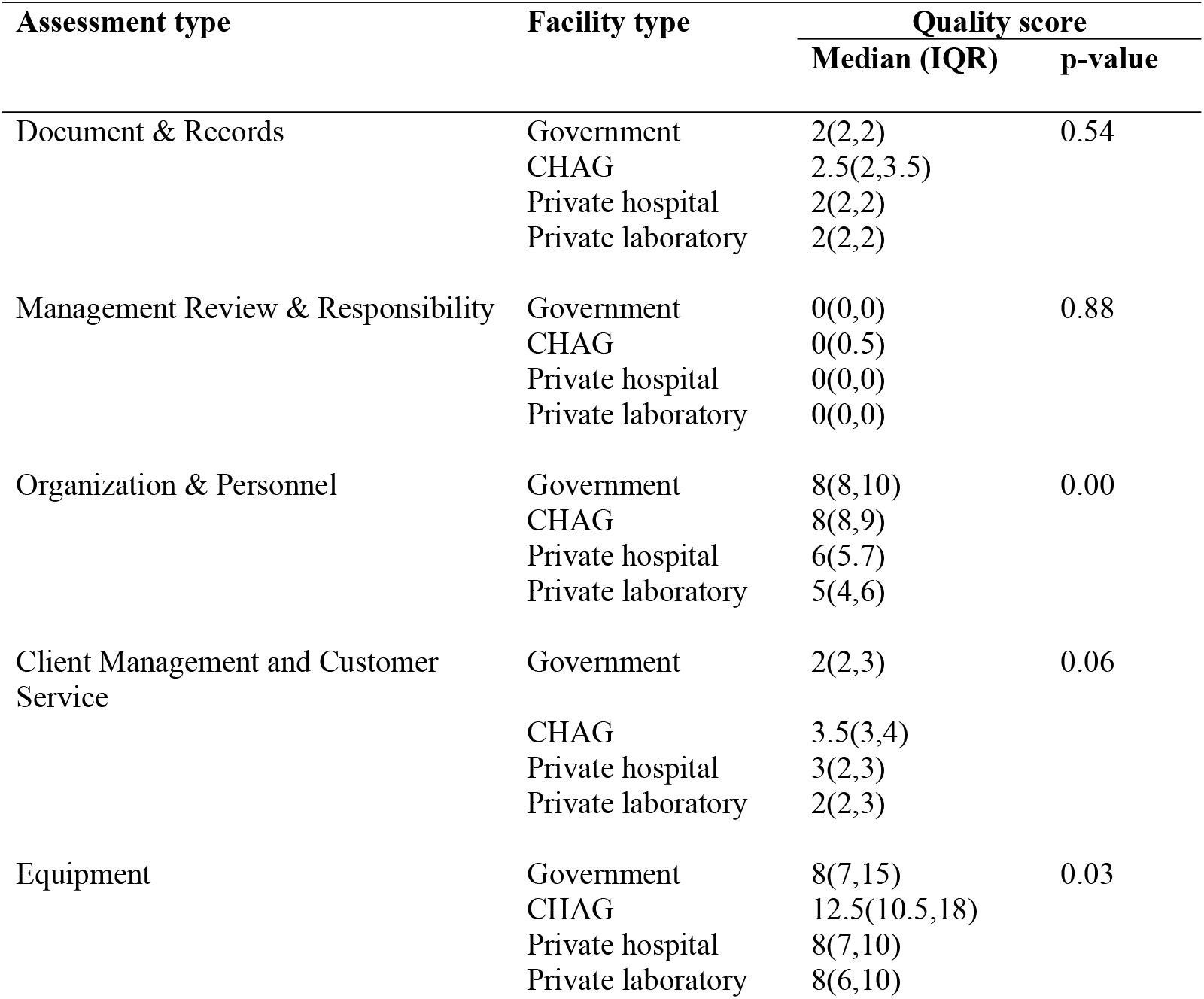
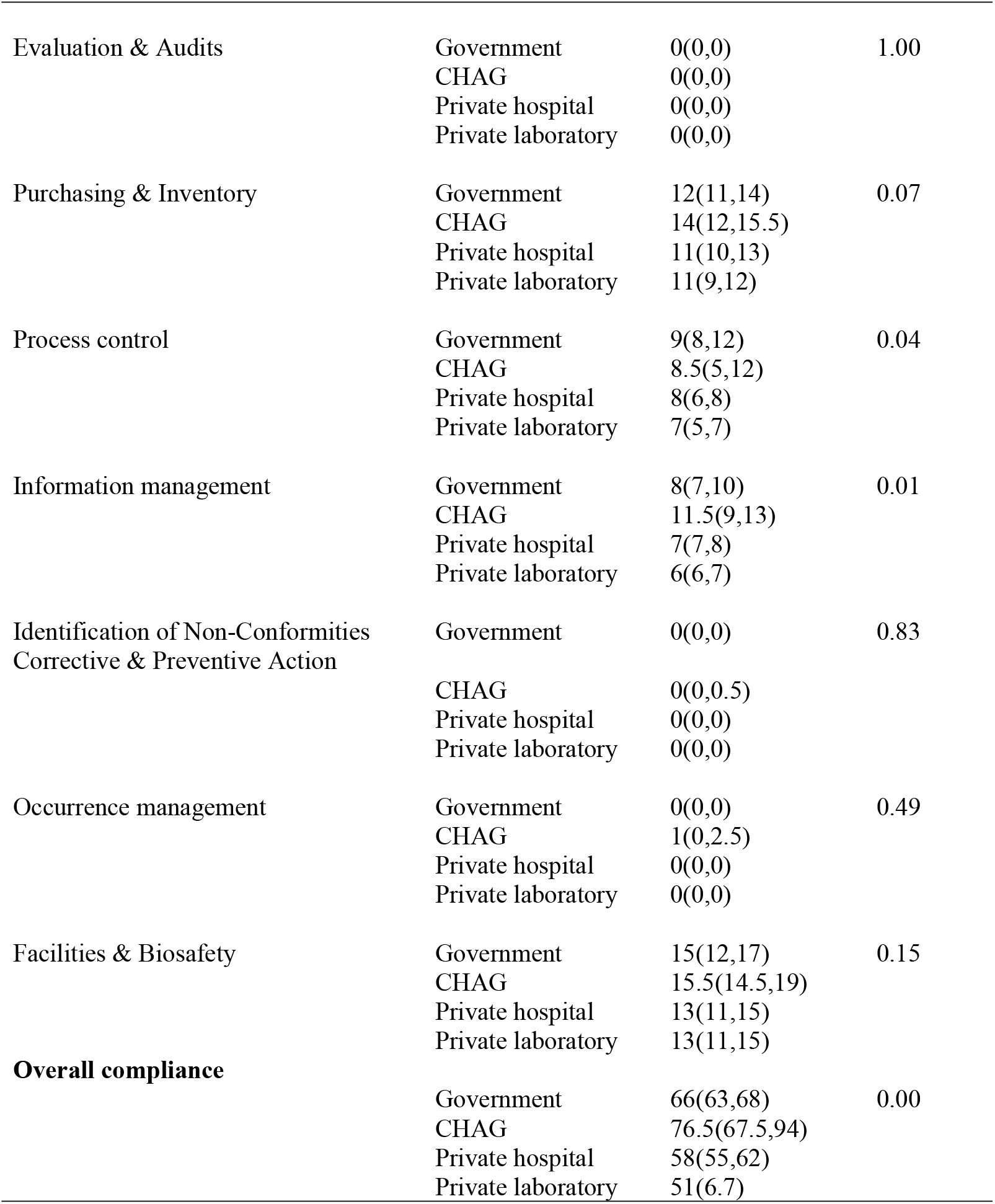
Differences in Laboratory Compliance Score by Facility Type.

### Differences in laboratory compliance score by facility type

Kruskal-Wallis Test was conducted to examine the differences in overall compliance scores and the quality system essential scores according to the types of health facilities. There were significant differences in the overall median compliance score (p=0.00) among the four (4) categories of health facilities. Similarly, there were significant differences in the following quality system essentials scores across facility types: Organisation & Personnel (p=0.00), Equipment (p=0.03), Process control (p=0.04) and information management (p=0.01).

## Discussion

The study showed that overall compliance with standard laboratory practice across all types of health facilities was low. None of the 43 medical laboratories assessed met or surpassed the minimum (150) standards for quality, as defined by the WHO SLIPTA checklist.

Although the level of compliance was low across facility types, government and CHAG facilities performed better than private health facilities. The low compliance with standards by laboratories seen in private health facilities could be attributed to inadequate resource flow to the private health sector and weak management and organizational capacity. Previous assessments of the private sector revealed that inadequate resources have resulted in poor infrastructure, obsolete equipment, inadequate human resource capacity and poor quality of care [12,13].

The low compliance observed across health facility types could be attributable to the fragmented regulation of laboratory services and the marginalization of laboratory services. The attitude of laboratory professionals towards quality could also be the reason for the poor performance of laboratories. The attitude of laboratory personnel has been identified as a barrier to providing quality laboratory services[14]. Kalra & Kopargaonkar (2016) further intimated that the quality components of a laboratory process are seen as “additional work” which interferes with their actual jobs by a workforce that is overworked and exhausted [14]. Hence, a change in attitude may be brought about by incorporating quality elements into the workplace culture and providing rewards for quality promotion actions and accomplishments.

It is interesting to note that Agboli et al. (2018), in their assessment of selected medical laboratories in healthcare facilities in the Volta region, Ghana, found that none of the laboratories met or surpassed the minimum compliance score, similar to the findings in this current study [10]. A recent survey conducted by Mulleta et al. (2021) in their assessment of laboratories in Ethiopia [15] found that nearly 80% (79.8%)of laboratories could not meet the minimum(<150) requirement for quality laboratory service provision, corroborating what was observed in this study[15]. Similarly, Mesganaw et al. (2023), in their systematic review of studies on medical laboratory quality management and challenges in Ethiopia, found that 91.7% of quality management practices were inadequate and the quality of medical laboratories was poor [16].

Furthermore, this study found that the performance of all laboratories in the twelve-quality system essentials was below standard. A recent assessment of laboratories in the Philippines made similar observations [17]. The quality system essentials are the building blocks of quality management systems and must be included in laboratory quality systems to ensure accurate, reliable and timely laboratory results. It is, therefore, not surprising that laboratories performed poorly in the compliance scores. The WHO indicate that although implementing quality management does not guarantee an error-free laboratory, it helps ensure high-quality laboratory services that detect and prevent errors from recurring [18]. It is, therefore, imperative that medical laboratories implement a quality management system to minimise errors and maintain high-quality laboratory services.

Inadequate quality has been identified as a barrier to providing optimal laboratory services in low and middle-income countries [19]. Medical laboratory services contribute about 70% of the data required for patient treatment and management, and therefore, quality laboratory services are a foundation for better patient management. The current finding suggests that medical laboratories are not fulfilling their mandate as required by standard practice and cannot play their crucial role in the healthcare delivery system due to poor compliance with standard laboratory practice. Medical laboratories operate in a complex environment, and ensuring high-quality services requires rigorous implementation of standards and guidelines [2].

The disproportionate distribution of laboratory professionals’ results from recruitment without recourse to the workload and expertise of laboratory professionals. Schneidman et al. (2014), in their assessment of neglected cadres in the healthcare system on the continent, indicate that, in sub-Saharan Africa, Laboratory professionals are predominantly among the neglected cadres in healthcare systems; often there are inadequate numbers, a skewed distribution, and limited career opportunities [20]. They further argued that insufficient professionals consequently affect the provision of quality laboratory service [20]. It is, therefore, not surprising that the current study made a similar observation eight years on; this is quite a worrying trend and needs to be addressed by health managers. Inadequate laboratory professionals affect service performance, as reflected in the low compliance scores recorded across all facility types in this study. Wilson and colleagues (2018), in their assessment of pathology and laboratory services in low and middle-income countries, identified inadequate human resource capacity as a critical barrier to providing medical laboratory services [19].

The lack of phlebotomists in the medical laboratory set-up, as evidenced in the current study, is a major concern. The study found that only tertiary health facilities had some phlebotomists; however, it was inadequate for the operation of the facility. The term “phlebotomy” refers to the procedure of drawing venous blood, and Phlebotomists are healthcare professionals who have received training to collect blood samples [21]. According to Ghana’s health sector staffing norm, only regional and tertiary hospitals must have a phlebotomist [22]. The lack of a framework for phlebotomy and its resultant lack of phlebotomists across the tiers of laboratory practice in Ghana’s health sector is a grave deficiency. The need for phlebotomists in the diagnostic process is highlighted because 60% of laboratory errors occur due to improper methods for collecting venous blood samples in the laboratory testing process. [23]. According to Waheed et al. (2013), even the most advanced laboratory equipment cannot provide accurate results from a specimen that has been improperly collected [24]. Hence, phlebotomists should be considered an essential component of the healthcare system, as they are important in providing suitable laboratory specimens for testing [21]. Mbah (2014) observed that employee cross-training for numerous duties is required in most healthcare facilities across Africa due to resource constraints, potentially eroding specialized skills and leading to excessive workloads (Mbah, 2014). However, in his assessment of phlebotomy and quality in the African laboratory, he argued that using skilled phlebotomy-specific laboratory personnel could significantly reduce pre-analytical error rates in the laboratory diagnostic value chain [25].

Therefore, there is an urgent need to institute pragmatic measures to address the quality gap and related factors that contribute to providing quality medical laboratory services in Africa, particularly in Ghana.

## Conclusion

None of the 43 medical laboratories assessed met or exceeded the minimum standards for quality. The study also revealed an uneven distribution of laboratory personnel across different facility types, and most laboratories lacked phlebotomists. Medical laboratories have been slow to adopt current quality management techniques to enhance the quality of laboratory services. The poor compliance of medical laboratories with basic quality standards is concerning and could significantly affect the quality of laboratory services.

## Data Availability

The data that support the findings of this study are available on request from the corresponding author, G.C.A. The data are not publicly available due to containing information that could compromise the privacy of health facilities.

## Acknowledgement

We would like to thank the research team, all hospitals, Health Directors, Health Service Administrators, Medical Superintendents, Nurse Managers, and Laboratory Managers who assisted with the recruitment of study participants and data collection.

